# Adjusting to Disrupted Assessments, Placements and Teaching (ADAPT): a snapshot of the early response by UK medical schools to COVID-19

**DOI:** 10.1101/2020.07.29.20163907

**Authors:** Anmol Arora, Georgios Solomou, Soham Bandyopadhyay, Julia Simons, Alex Osborne, Ioannis Georgiou, Catherine Dominic, Shumail Mahmood, Shreya Badhrinarayanan, Syed Rayyan Ahmed, Jack Wellington, Omar Kouli, Robin Jacob Borchert, Joshua Feyi-Waboso, Scott Dickson, Savraj Kalsi, Dimitrios Karponis, Timothy Boardman, Harmani Kaur Daler, Abbey Boyle, Jessica Speller, Conor S Gillespie, Jie Man Low, Ratnaraj Vaidya, Ngan Hong Ta, Steven Aldridge, Jonathan Coll Martin, Natasha Douglas, Mary Goble, Tayyib Abdel-Hafiz Goolamallee, Emma Jane Norton, Andre Chu, Inshal Imtiaz, Oliver P Devine

## Abstract

**Background:** Medical school assessments, clinical placements and teaching have been disrupted by the COVID-19 pandemic. The ADAPT consortium was formed to document and analyse the effects of the pandemic on medical education in the United Kingdom (UK), with the aim of capturing current and future snapshots of disruption to inform trends in the future performance of cohorts graduating during COVID-19.

**Methods:** Members of the consortium were recruited from various national medical student groups to ensure representation from medical schools across the UK. The groups involved were: Faculty of Medical Leadership and Management Medical Students Group (FMLM MSG); Neurology and Neurosurgery Interest Group (NANSIG); Doctors Association UK (DAUK); Royal Society of Medicine (RSM) Student Members Group and Medical Student Investigators Collaborative (MSICo.org). In total, 29 medical schools are represented by the consortium. Our members reported teaching postponement, examination status, alternative teaching provision, elective status and UK Foundation Programme Office (UKFPO) educational performance measure (EPM) ranking criteria relevant to their medical school during a data collection window (1st April 14:00 to 2nd April 23:59).

**Results:** All 29 medical schools began postponement of teaching between the 11th and 17th of March 2020. Changes to assessments were highly variable. Final year examinations had largely been completed before the onset of COVID-19. Of 226 exam sittings between Year 1 and Year 4 across 29 schools: 93 (41%) were cancelled completely; 14 (6%) had elements cancelled; 57 (25%) moved their exam sitting online. 23 exam sittings (10%) were postponed to a future date. 36% of cohorts with cancelled exams and 74% of cohorts with online exams were granted automatic progression to the next academic year. There exist 19 cohorts at 9 medical schools where all examinations (written and practical) were initially cancelled and automatic progression was granted.

**Conclusions:** The approaches taken by medical schools have differed substantially, though there has been universal disruption to teaching and assessments. The data presented in this study represent initial responses, which are likely to evolve over time. In particular, the status of future elective cancellations and UK Foundation Programme Office (UKFPO) educational performance measure (EPM) decile calculations remains unclear. The long-term implications of the heterogeneous disruption to medical education remains an area of active research. Differences in specialty recruitment and performance on future postgraduate examinations may be affected and will be a focus of future phases of the ADAPT Study.

## Background

The COVID-19 pandemic continues to have a pervasive impact on healthcare around the world. In addition to a dramatic restructuring of healthcare systems, education for the next generation of doctors has needed to rapidly adapt to a complex and changing situation. Teaching, both in the classroom and at the bedside, became virtually impossible as university campuses closed, and teaching hospitals prioritised essential clinical work. Though the situation has improved at the time of writing, the medical community is braced for further waves of disease in the coming Winter months.

In the UK, medical schools operate within a regulatory framework that enables significant autonomy, providing certain minimum outcomes are met by the point of graduation^1^. So long as schools meet standards set by the GMC, they broadly determine their own curriculum; semester dates; and the amount of assessment and teaching that takes place. As a result, heterogeneity exists in student experience between UK medical schools. The impact of these differences has been the topic of much previous research by ourselves and others^2–5^.

Alterations to assessment and teaching in response to COVID-19 provide a rare opportunity to measure the impact of changes to the way medical students learn and are assessed. It is unlikely these changes would have otherwise occurred with such rapidity or magnitude. In order to track the impact of such changes, we convened a collaborative group of student researchers at each UK medical school and formed the ‘Adjusting to Disrupted Assessments, Placements and Teaching’ or ‘ADAPT’ Consortium. Here, we report an early snapshot of our multi-phase project by describing the response of 29 UK medical schools in terms of changes to assessment, clinical placement and teaching provision between March and early April 2020.

## Method

The ADAPT Consortium comprises 34 student members and was populated by students representing each of the following medical student bodies: Faculty of Medical Leadership and Management Medical Students Group (FMLM MSG); Neurology and Neurosurgery Interest Group (NANSIG); Doctors Association UK (DAUK); Royal Society of Medicine (RSM) Student Members Group; or Medical Student Investigators Collaborative (MSICo.org). The contributors to the ADAPT consortium are listed in Appendix A.

A data collection matrix was piloted and finalised (Appendix B), collecting data on the following metrics: teaching postponement, examination status, alternative teaching provision, elective status and ranking information. For each year group, students were asked to record dates teaching was postponed from, as well as anticipated dates for the resumption of placements. We also recorded the status of examinations and teaching activities, both written and practical. Data were collected for every year at each medical school in order to capture variation between years; since it has been noted that the type of teaching received differs along progression through medical school^4^.

The data matrix was piloted with a small number of consortium members. An agreed data collection window started on the 1st April 2020 at 14:00hrs GMT and ended on the 2nd April 23:59hrs GMT. Given rapidly changing local response, the collection window was deliberately narrow to ensure comparability between schools.

In order to determine the effect of COVID-19-related disruption on planned summative examinations for the 2019-2020 academic year, consortium members reported on the status of examinations on a discrete scale which included: ‘all cancelled’; ‘elements cancelled’; ‘elements moved online; ‘all examinations moved online’; ‘all examinations postponed’; ‘exams to take place as planned’; ‘changes to examinations undecided’; and ‘examinations already completed prior to COVID-19-related disruption’. If a particular type of examination was not ordinarily performed for a particular year group, this was reported as ‘not applicable’. Students were also asked to report information regarding: elective status, educational performance measure (EPM) calculation status, progression details and teaching status for each year.

The research presented in this manuscript, which is the initial phase of the ADAPT Study, was determined to be exempt from ethical review by the University College London (UCL) Research Ethics Committee (30/03/20).

## Results

### Effect of COVID-19 on placements and teaching

On the 13th of March 2020, the Medical Schools Council (MSC) released a statement stating that:

~~~
*“*…*where prioritisation is needed final year students should be prioritised” and that “*…*placements can be paused and equivalent experience provided later on in the course”* ^*6*^
~~~

Whilst one medical school had already postponed some teaching, the MSC statement was followed by the rapid postponement of at least some teaching at 11 of 29 schools on the same day. Over the subsequent 4 days, all medical schools postponed teaching to some degree. On the 18th of March, the UK government announced national closures of schools and universities. Taken together, these findings suggest a rapid and coordinated effort to minimise mutual exposure of students to patients in the clinical environment which was completed by mid-March. From this point onwards ‘normal’ teaching did not take place at any medical school included in this study.

### Effect of COVID-19 on planned summative assessments

One of the core, fixed activities of medical schools is the provision of rigorous assessments to assure the quality of their graduates. The distribution of planned changes to assessments, as recorded in April 2020, is shown in **Figure 2A**. Due to the earlier assessment period typically designated for final year examinations, a large number of schools (12 of 29) had already completed all forms of assessment for their final year students prior to the disruption caused by COVID-19. A further 4 schools had already completed their final year practical assessments. This meant that at the majority of schools, written finals examinations were completed either as planned (16/29) or using modified online assessments (8/29). In the earlier years (Years 1-4), only 5 out of 232 exam sittings had been completed nationally by the time of our study. Of the 226 remaining: 93 (41%) sittings were cancelled completely; 14 (6%) had elements cancelled; 57 (25%) moved their exam sittings online. 23 exam sittings (10%) were postponed to a future date (undeterminable by our study). Over the entire length of each course at each school, 9% of exam sittings were taking place as planned or had decisions pending regarding their status at the time of our data collection.

**Figure 1.**
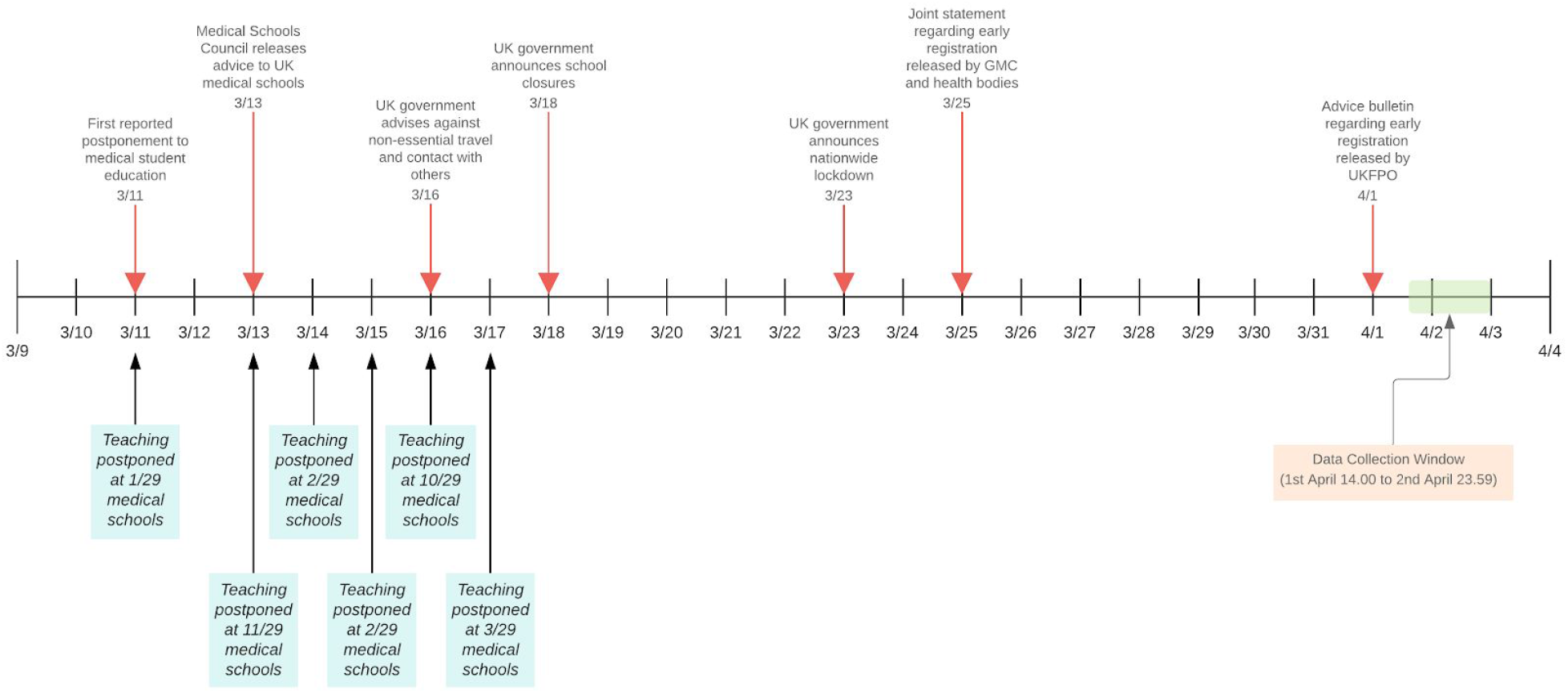
Timeline of actions taken by medical schools between 09/03/20 and 03/04/20. *Red arrows* indicate key policy decisions made by government and regulatory bodies. *Blue boxes* indicate progressive postponement of teaching at UK medical schools. *Orange box* indicates the ADAPT Study data collection window.

**Figure 2.**
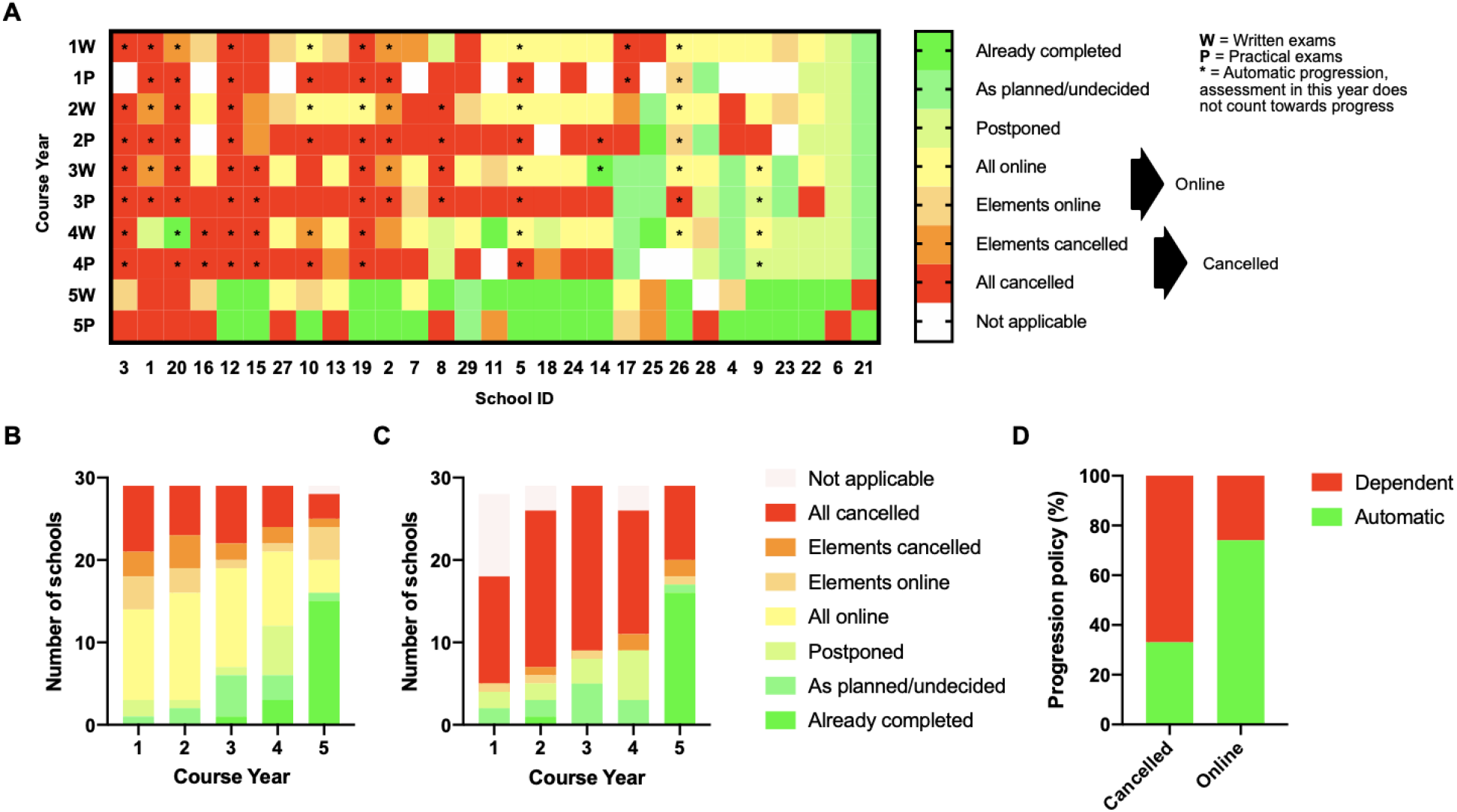
Effect of COVID-19-related disruption on summative assessment at UK medical schools. Data were collected on the status of summative written (W) and practical (P) examinations for years 1-5 at 29 UK medical schools. **(A)** Heatmap showing the status of examinations at each school, sorted by column total from schools with predominantly cancelled examinations (left) to schools with completed or unchanged examinations (right). Asterisk (*) denotes year groups with automatic progression; where the results of assessment would not count towards progression to the next academic year. Stacked bar charts show the distribution of exam status by course year for both written **(B)** and practical **(C)** examinations. **(D)** Progression policies for cohorts where assessment was confirmed as having been cancelled (entirely or partially) or moved online (entirely or partially) represented as a proportion of automatic (green) versus exam-dependent (red) progression.

**Figure 3.**
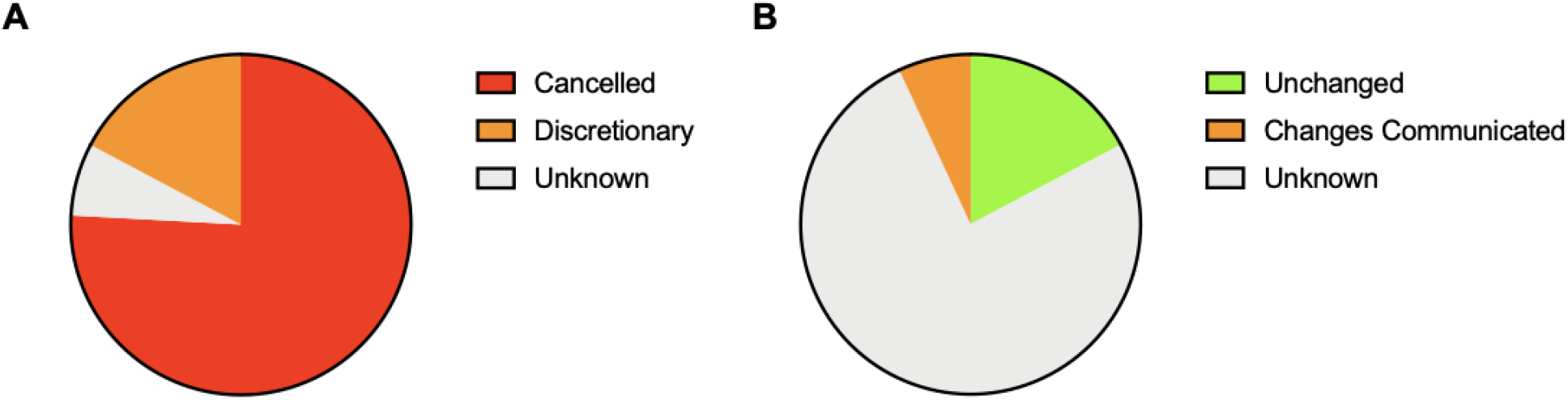
Known changes to electives **(A)** and educational performance measure (EPM) calculations **(B)** as a result of COVID-19.

Though cancellation was the most common outcome overall, there were considerable differences between written and practical examinations. Whilst 37 of 116 (32%) of written exam sittings were cancelled across Years 1-4 **(Figure 2B, red bars)**; this was increased to 70 of 99 (70%) for practical exam sittings **(Figure 2C, red bars)**. The practical exam sittings that were not cancelled were mostly either postponed until the next academic year (13 of 99 (13%)) or undecided at the time of our study (12 of 99 (12%)). Of the written exam sittings not cancelled, 45 of 116 (39%) were scheduled to take place online; 10 of 116 (9%) were postponed; with the remainder already completed or with decisions pending. These findings highlight the challenges of conducting practical examinations under the conditions brought about by the spread of COVID-19 anticipated in March 2020.

Significant disruption to assessment and teaching undoubtedly influenced progression decisions for the 2019-2020 academic year, resulting in a large number of schools introducing automatic progression for their cohorts (**Figure 2A**, asterisks). When all, or only elements of a particular exam diet were cancelled, 36% of cohorts were given automatic progression (**Figure 2D**). This may seem low, however, it is possible that schools cancelling exams after the COVID-19 outbreak were more likely to do so if students had already fulfilled sufficient progression criteria (e.g. continuous assessment or modular assessment strategies). Notably, when planned exams had been moved online, 74% of those cohorts were granted automatic progression (**Figure 2D**).

### Changes to electives and calculation of the education performance measure (EPM) as a result of COVID-19

#### Changes to the elective

All UK medical students undertake a period of elective study during the final or penultimate year of their course. A large proportion of these study periods are spent experiencing healthcare in international settings^7^. Global travel restrictions put in place prior to and during March 2020 created significant pressure on medical schools to form policy positions on this matter ^8,9^. On the 17th of March, the UK Foreign and Commonwealth Office (FCO) issued a statement advising against all non-essential foreign travel, though many elective destinations had already imposed travel restrictions for arrivals. By the time of our data collection window, 22 of 29 (76%) medical schools had officially cancelled their elective for the 2019-2020 academic year; 5 of 29 (17%) had made it discretionary; with the remaining 2 schools not having made a decision.

#### Changes to the EPM

Another key part of the sequence of events taking place in the final year of medical school is the calculation of the education performance measure or ‘EPM’. This is determined differently at each school but typically consists of the ranking of students into decile bands according to performance in examinations over two or more years of medical school^10^. Given that many assessments whose results are used to determine the EPM were expected to be cancelled or conducted in a different format, we speculated that schools may have given students advanced notice of any changes to calculation methods. This was largely not the case by early April; with 22 of 29 (76%) of schools reporting no announcement of changes. 5 of 29 (17%) schools reported that the EPM calculation would not change. The 2 schools that had communicated EPM changes told that the measure would give an increased weighting to the previous years’ exams, in both cases this was year 3 exams. One of the medical schools who reported that their EPM calculation method would remain the same gave the caveat that for final years only, the postponed exams would not be included.

## Discussion

The disruption caused by COVID-19 to medical education has been swift and substantial. The ADAPT Study represents the first comprehensive description of the early national response. Students were promptly removed from classroom and clinical learning environments across the country over a short period of time. This response is likely attributable to national coordination by the Medical Schools Council (MSC), with evidence of a temporal association between the MSC statement on 13th of March and the subsequent postponement of teaching at all schools over the next few days. This initial report is unable to address how long teaching postponement is expected to last. Anecdotally, our authors report a consensus of “Autumn 2020” specified by the majority of schools, which corresponds with a statement from the MSC on the 20th of May:

~~~
*“*…*we anticipate that most medical schools will be able to restart clinical placements in September 2020”*^*11*^
~~~

The performance and preparedness of graduates varies between UK medical schools^2,5^. This is related to the volume of assessment^3^; as well as the types and subjects of teaching experienced during medical school^5^. The *amount* of formal teaching a student is exposed to varies significantly between medical schools^4^, but has not been found to play a major role in later postgraduate performance^4^. Variation in the date of return between UK schools, and variation in the teaching provisions that have taken place during the postponement of teaching, is therefore of considerable interest for future investigations that seek to determine the impact of this disruption to later professional performance. Previous studies have documented the exact composition of UK medical courses^4,5^. The unique circumstances of students in the ‘COVID-19 cohorts’ provides an opportunity to determine the impact of these changes to teaching on later performance and career destinations. The evaluation of precisely *what* teaching has been reduced, and what teaching has been provided in place of ‘normal’ scheduling is best determined in retrospect, and as such will form the basis for future iterations of the ADAPT Study.

Whilst it is clear that students were stood down from classroom and clinical environments in a coordinated manner over the space of several days, changes to assessment were highly variable and formed the main focus of the present phase of our study. Only a small fraction (9%) of assessments were scheduled to go ahead as planned at the time of our data collection. This suggested that the majority of schools had made decisions about their assessments by the time of our data collection period at the start of April. Due to the nature of medical school finals (typically in Year 5), a large proportion of final year exams had been conducted prior to the disruption caused by COVID-19. This was undoubtedly helpful in meeting the criteria outlined by the General Medical Council for the provisional registration of final year medical students taking up Foundation Interim Year 1 (FiY1) jobs in UK hospitals at the peak of this initial wave of the pandemic^12^. Much of our analysis, therefore, focussed on Years 1-4. Just under half of all exam sittings were cancelled, with approximately a quarter moved online. The remainder were undecided, planned to postpone, or in some cases were already complete. This will result in cohorts graduating with less exposure to both written and practical assessments as well as providing an opportunity to understand the relationship between undergraduate assessment volume and performance in postgraduate examinations ^3,5^.

Mirroring the shift towards telemedicine in clinical practice, there has been a rapid move to introduce online assessments in place of traditional written examinations. 39% of written examinations were planned to take place in this format. We found that when assessments were cancelled, approximately one-third of cohorts were granted automatic progression. This left a substantial proportion of cohorts with cancelled exams, and progression determined by other methods. There was not sufficient granularity in our data collection to identify how progression was calculated, but anecdotally, the authors report that estimates based on performance in previous years examinations or satisfactory completion of logbooks were used in some cases. We identified 19 cohorts at 9 medical schools where all examinations (written and practical) were cancelled and automatic progression was granted.

The initial phase of our study did not capture the precise nature of how online assessments are planned to be conducted. In the next phase of the study we will identify further details pertaining to: delivery methods (including the software used); provisions for invigilation (including online proctoring); duration and pace of the assessment; and standard setting. Furthermore some decisions surrounding assessments have likely evolved since our initial data collection period and will be reviewed in future iterations of the study.

Preliminary data pertaining to decisions about EPM calculations for the 2020-2021 academic year were included to give an indication of the status at the time of our data collection. It is clear that these areas continue to change also, and at the time of data collection, the majority of schools had not communicated any changes to their students. Anecdotally, the authors report that this has now changed at some schools. Further data will be collected in the next phase of the study to determine the nature of these changes. Crucially, we aim to understand how EPM calculations for the 2020-2021 academic year will differ from the previous year: including whether grading from online assessments; assessments not used for progression; scoring of logbooks; and grade estimation will be used.

## Conclusions

This study represents the first comprehensive quantitative snapshot of the early disruption caused by COVID-19 to undergraduate medical education in the UK. At the time of writing, normal teaching has been suspended at all UK medical schools. Our findings show heterogeneity in how schools have altered their assessment in response to the pandemic; with exams broadly divided between being cancelled or moved online. A substantial minority of schools plan to progress students to the next academic year without sitting exams.

The long term goal of the ADAPT Study remains to investigate the impact of the disruptions caused by COVID-19 on postgraduate performance of UK medical graduates. In the next phase of this study, we plan to map the reintroduction of normal teaching programmes at the start of the 2020-2021 academic year; report additional changes to teaching and examinations enacted by medical schools; quantify the reduction in teaching and assessment over the duration of the lockdown; and compare changes to EPM calculation between schools. We also intend to measure whether the decisions made in the early response are sustained or altered as the pandemic evolves. We may begin to see convergence of decisions as placements begin to resume or divergence if regional lockdowns begin to disturb local medical schools.

## Data Availability

The authors declare that the key aggregated data supporting the findings of this study are available within the article (Figure 2). Other raw data are available from the corresponding author, OPD, upon reasonable request.

## Ethics declaration

None of the data presented in this manuscript are personal data or identifiable to individual medical schools. The work presented was exempted from ethical review by the University College London (UCL) Research Ethics Committee (30/03/20).

### Appendix A

#### Associated Student Organisations

Faculty of Medical Leadership and Management Medical Students Group (FMLM MSG); Neurology and Neurosurgery Interest Group (NANSIG); Doctors Association UK (DAUK); Royal Society of Medicine (RSM) Student Members Group; or Medical Student Investigators Collaborative (MSICo.org)

#### ADAPT Study Lead

Oliver P Devine

#### ADAPT Steering Group

Oliver P Devine (FMLM MSG, MSICo.org), Anmol Arora (FMLM MSG), Georgios Solomou (FMLM MSG, NANSIG), Soham Bandyopadhyay (NANSIG), Julia Simons (DAUK), Alex Osborne (RSM)

#### ADAPT Senior Writing Group

Anmol Arora, Georgios Solomou, Oliver P Devine

#### ADAPT Data Collectors

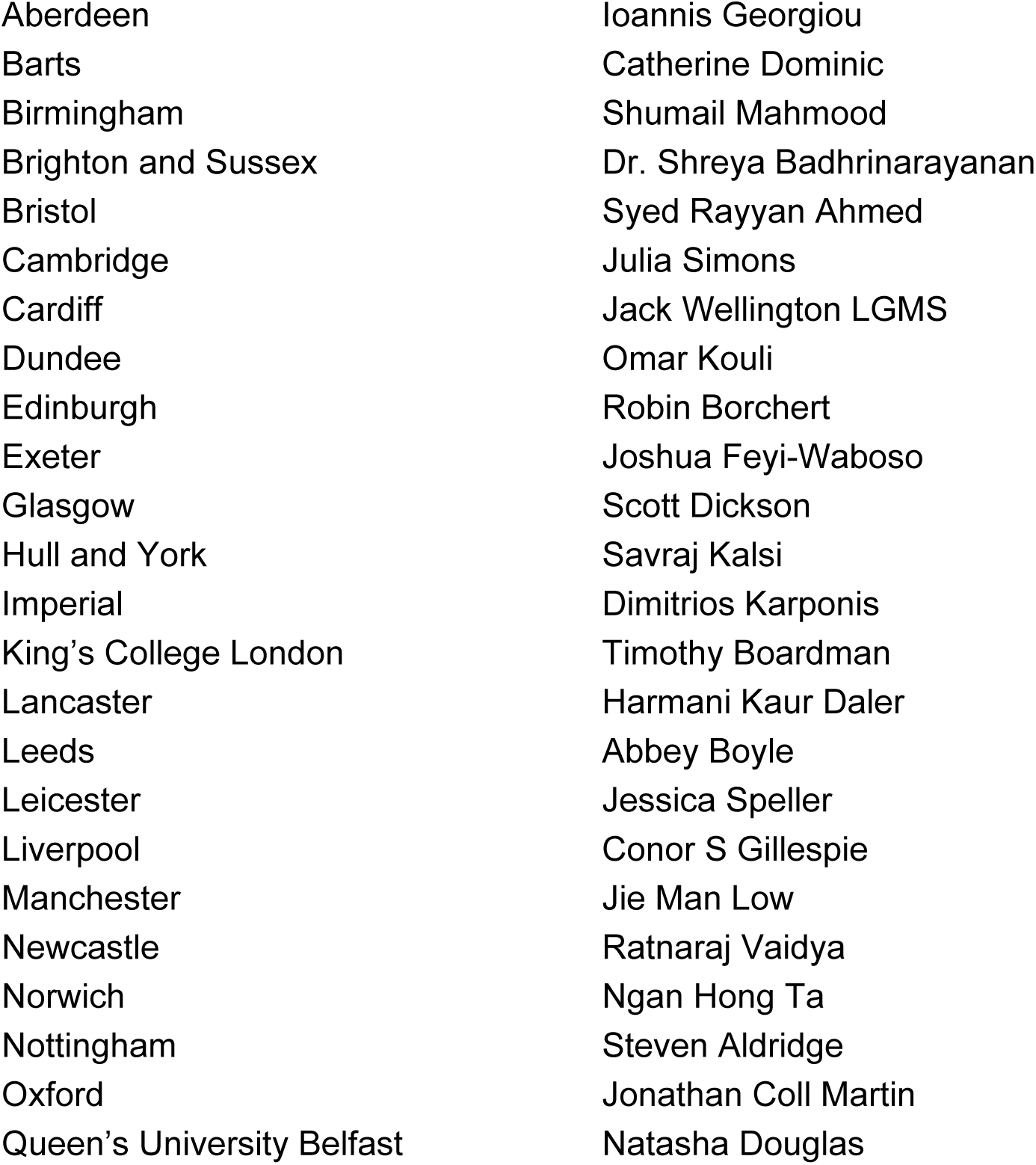
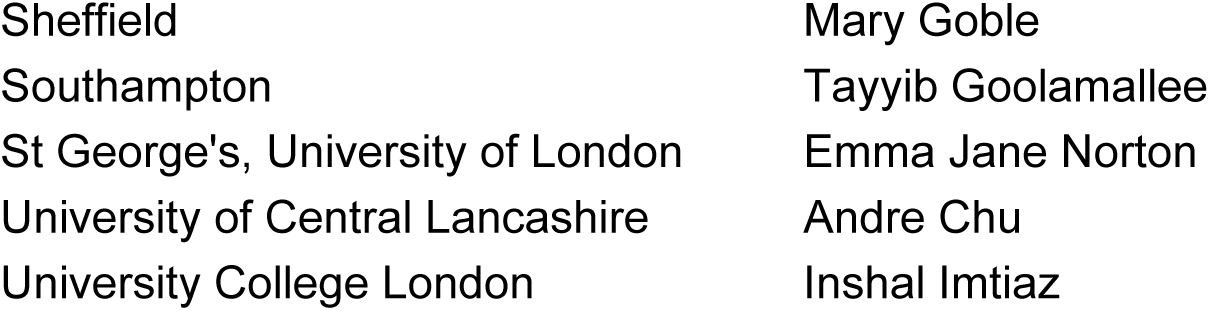

### Appendix B

#### Year 1

*Classroom teaching postponed?*

*Postponement start date*

*Postponement end date (current plan)*

*Clinical teaching postponed?*

*Postponement start date Postponement end date (current plan)*

*Alternative teaching provision in place*

*Alternative teaching details (free text)*

*Written assessment status*

*Practical assessment status*

*Further details about assessments (free text)*

*Progression (free text)*

#### Year 2

*Classroom teaching postponed?*

*Postponement start date*

*Postponement end date (current plan)*

*Clinical teaching postponed?*

*Postponement start date Postponement end date (current plan)*

*Alternative teaching provision in place Alternative teaching details (free text)*

*Written assessment status*

*Practical assessment status*

*Further details about assessments (free text)*

*Progression (free text)*

#### Year 3 (1st clinical)

*Classroom teaching postponed?*

*Postponement start date*

*Postponement end date (current plan)*

*Clinical teaching postponed?*

*Postponement start date*

*Postponement end date (current plan)*

*Alternative teaching provision in place*

*Alternative teaching details (free text)*

*Written assessment status*

*Practical assessment status*

*Further details about assessments (free text)*

*Progression (free text)*

#### Year 4 (2nd clinical)

*Classroom teaching postponed?*

*Postponement start date*

*Postponement end date (current plan)*

*Clinical teaching postponed?*

*Postponement start date*

*Postponement end date (current plan)*

*Alternative teaching provision in place*

*Alternative teaching details (free text)*

*Written assessment status*

*Practical assessment status*

*Further details about assessments (free text)*

*Progression (free text)*

#### Year 5 (final year)

*Classroom teaching postponed?*

*Postponement start date*

*Postponement end date (current plan)*

*Clinical teaching postponed?*

*Postponement start date*

*Postponement end date (current plan)*

*Alternative teaching provision in place*

*Alternative teaching details (free text)*

*Written assessment status*

*Practical assessment status*

*Further details about assessments (free text)*

*Progression (free text)*

#### Compulsory intercalated year

(If intercalation is not compulsory please leave this section blank)

*Classroom teaching postponed?*

*Postponement start date*

*Postponement end date (current plan)*

*Clinical teaching postponed?*

*Postponement start date*

*Postponement end date (current plan)*

*Alternative teaching provision in place*

*Alternative teaching details (free text)*

*Written assessment status*

*Practical assessment status*

*Further details about assessments (free text)*

*Progression (free text)*

#### General

*Have online, at-home, summative exams ever been performed at your school before?*

#### Electives

*What is the current status of electives? (free text)*

#### EPM

*Have there been any announcements regarding the calculation of the EPM? (free text)*

#### Other

*Are there any other details you wish to report?*

